# Deep learning of SARS-CoV-2 outbreak phylodynamics with contact tracing data

**DOI:** 10.1101/2024.06.10.24308687

**Authors:** Ruopeng Xie, Dillon C. Adam, Shu Hu, Benjamin J. Cowling, Olivier Gascuel, Anna Zhukova, Vijaykrishna Dhanasekaran

## Abstract

Deep learning has emerged as a powerful tool for phylodynamic analysis, addressing common computational limitations affecting existing methods. However, notable disparities exist between simulated phylogenetic trees used for training existing deep learning models and those derived from real-world sequence data, necessitating a thorough examination of their practicality. We conducted a comprehensive evaluation of model performance by assessing an existing deep learning inference tool for phylodynamics, PhyloDeep, against realistic phylogenetic trees characterized from SARS-CoV-2. Our study reveals the poor predictive accuracy of PhyloDeep models trained on simulated trees when applied to realistic data. Conversely, models trained on realistic trees demonstrate improved predictions, despite not being infallible, especially in scenarios where superspreading dynamics are challenging to capture accurately. Consequently, we find markedly improved performance through the integration of minimal contact tracing data. Applying this approach to a sample of SARS-CoV-2 sequences partially matched to contact tracing from Hong Kong yields informative estimates of SARS-CoV-2 superspreading potential beyond the scope of contact tracing data alone. Our findings demonstrate the potential for enhancing deep learning phylodynamic models processing low resolution trees through complementary data integration, ultimately increasing the precision of epidemiological predictions crucial for public health decision making and outbreak control.

## Introduction

Phylogenetic analysis of genomic sequence data offers a powerful toolkit for understanding the emergence, spread, and evolution of infectious diseases. As an interdisciplinary field, phylodynamics aims to integrate genomic and epidemiological data in a unified framework to extract detailed insights into epidemic history (Drummond et al., 2005; Stadler et al., 2013; Volz et al., 2009), population dynamics (Stadler & Bonhoeffer, 2013; Volz et al., 2009), and disease emergence (Pekar et al., 2022; Worobey et al., 2014). Its key advantage lies in providing independent information regarding epidemic history, complementing traditional epidemiological surveillance data (Vaughan et al., 2024; Voznica et al., 2022). This makes it invaluable for validating and substantiating findings from epidemiological modelling, particularly in contexts where conventional surveillance data are scarce and genomic sampling is randomized.

However, many conventional phylodynamic models based on likelihood approaches (e.g. maximum likelihood estimation and Bayesian approaches) are computationally intensive and can become practically unfeasible as the number of taxa increases (Hohna & Drummond, 2012). Addressing this issue sometimes involves likelihood-free methods such as approximate Bayesian computation (ABC) (Saulnier et al., 2017), which sidestep the need for direct likelihood calculations. More recently, deep learning methods such as PhyloDeep (Voznica et al., 2022) have emerged as another potential solution, enabling rapid estimation of epidemiological parameters from large phylogenetic trees in a matter of seconds. To achieve this, PhyloDeep utilizes deep neural network models trained against phylogenies simulated under well-established birth-death models: the basic birth-death model (BD) (Leventhal et al., 2014; Stadler et al., 2012), the birth-death model with exposed and infectious classes (BDEI) (Kuhnert et al., 2016; Stadler et al., 2013), and the birth-death model with superspreading (BDSS) (Stadler et al., 2013). PhyloDeep has also been validated for diversification analyses (Lambert et al., 2023) and viral phylogeography (Thompson et al., 2024).

Despite these methodological advancements, there are often discrepancies between the idealised phylogenetic trees, simulated from birth-death models, and real-world trees constructed from samples of empirical sequence data. This challenge is particularly pronounced for viral sequences arising from epidemics or outbreaks, which frequently yield many identical sequences, resulting in low resolution phylogenies with numerous polytomies. Examples include SARS-CoV-2, Mpox (monkeypox) virus (Paredes et al., 2024), and Respiratory syncytial virus (RSV) (Eden et al., 2022). As such, the implications of employing realistic phylogenetic trees for predictions using neural network models, trained on simulated or “ideal” trees, remain uncertain.

In this study, we utilize the PhyloDeep framework, employing the SARS-CoV-2 as a virus outbreak characterized by the BDSS model. Our analysis reveals that neural network models trained on “ideal” trees struggle to precisely predict epidemiological parameters, particularly those associated with superspreading events, when applied to realistic phylogenetic trees. Furthermore, we observe a notable enhancement in predictive accuracy upon integrating contact tracing data into realistic phylogenetic trees, thereby aligning them more closely with the “ideal” trees. We illustrate these findings using real SARS-CoV-2 data collected during the third and fourth waves of the epidemic in Hong Kong.

## Results

### Simulations of phylogenetic trees

Initially, we simulated 200,000 time-scaled trees using the BDSS model (Fig. 1, baseline tree). These trees serve as our reference “ideal” trees and capture transmission events at internal nodes consistent with the PhyloDeep framework. To emulate realistic SARS-CoV-2 phylogenetic trees, all baseline trees were transformed into genetic distance trees (Fig. 1, genetic baseline tree).This transformation relied on a binomial distribution of mutation counts given a mean substitution rate of 8×10^−4^ per site per year (see methods for details). Branches with lengths representing zero mutation were collapsed, resulting in trees with polytomies (Fig. 1, genetic polytomous tree), which were then randomly resolved using a coalescent approach, yielding binary trees (Fig. 1, genetic resolved tree). The number and size of polytomies in our simulated trees varied from 1 to 170 and 3 to 934, respectively, with a total tip range of 200 to 1000, encompassing those observed in SARS-CoV-2 trees in Hong Kong (Supplementary Figure S2). Lastly, each of the three transformed genetic distance trees were dated using LSD2 (To et al., 2016) (Fig. 1, dated baseline tree, dated polytomous tree, dated resolved tree). The latter four types of trees, including Genetic Polytomous Trees, Genetic Resolved Trees, Dated Polytomous, and Dated Resolved Trees, represent entirely altered topologies and are deemed realistic trees, as they can be generated from sequencing data using established software such as RAxML-NG (Kozlov et al., 2019), IQ-TREE (Nguyen et al., 2015), FastTree (Price et al., 2010) or TreeTime (Sagulenko et al., 2018). In contrast, the remaining three types, including Baseline Trees, Genetic Baseline Trees, and Dated Baseline Trees, retain a known correct topology that cannot be derived from sequence data alone (Fig. 1).

**Fig. 1.**
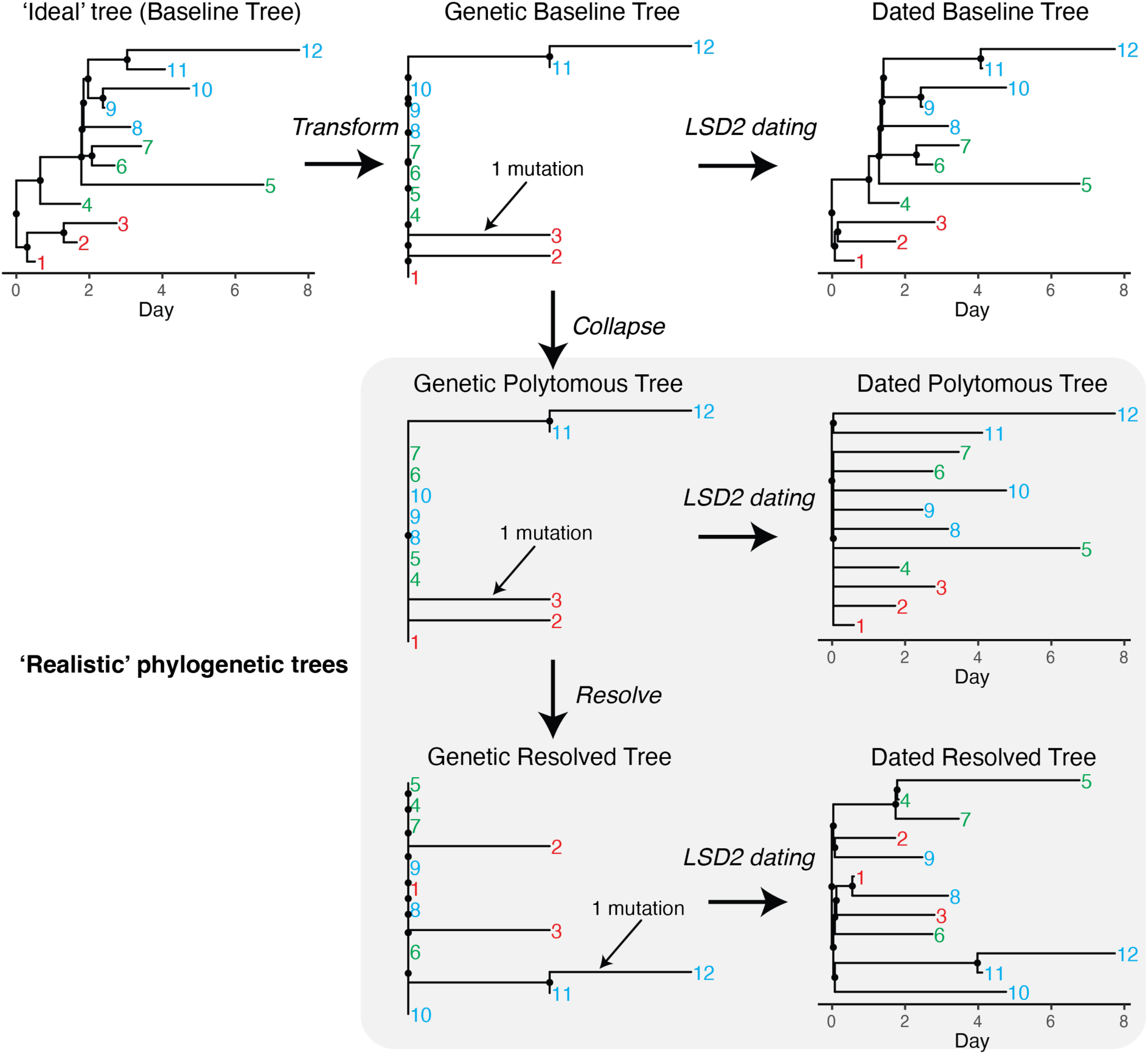
Examples of seven types of phylogenetic trees used in simulations. Internal nodes are marked as black dots, while tips are denoted by numerical labels. Among these, four trees represent realistic phylogenetic structures that can be derived from sequence data and are highlighted with a grey background. To effectively highlight the differences between realistic trees, which can be constructed from sequence data, and unrealistic trees, which cannot, tips have been color-coded into three distinct clusters.

### Performance comparison of neural network models for each type of phylogenetic tree

We utilized a dataset totalling 199,000 trees to train the neural network models, reserving 1,000 trees for validation purposes. Ensuring consistency across the models, we utilized identical summary statistics (SSs) representation and feed-forward neural networks (FFNNs) for each tree type, as used in PhyloDeep (Fig. 2). Specifically, for the three types of genetic distance trees, including Genetic Baseline Trees, Genetic Polytomous Trees and Genetic Resolved Trees, we adapted the 99 SSs designed for time-scaled trees to 90 SSs for genetic distance trees (refer to the Methods section). Consequently, we developed seven neural network models: Baseline-Model, Dated Baseline-Model, Dated Resolved-Model, Dated Polytomous-Model, Genetic Baseline-Model, Genetic Resolved-Model, and Genetic Polytomous-Model.

**Fig. 2.**
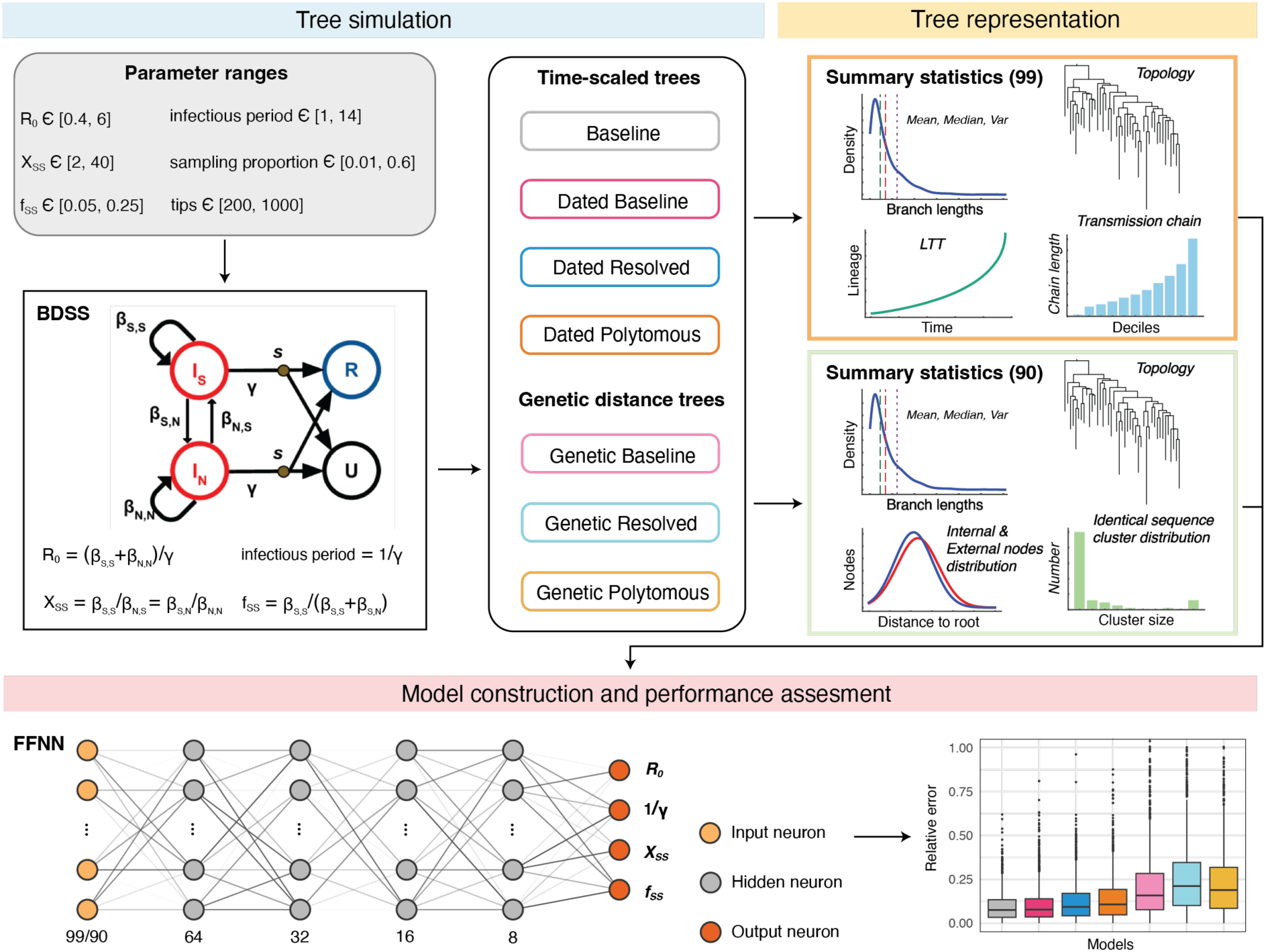
**An overview of training neural network models based on simulated phylogenetic trees.**

Our results show that models trained and tested on trees with unchanged topologies (i.e. Baseline-Model, Dated Baseline-Model, and Genetic Baseline-Model) did well in predicting all parameters. Estimates for *R_0_* and infectious period tended to exhibit greater accuracy compared to superspreading parameters (*X_ss_* and *f_ss_*) (Fig. 3A and Supplementary Table S1), which is consistent with the findings from PhyloDeep (Voznica et al., 2022). The Baseline-Model demonstrated superior performance with mean relative errors of 0.095 for *R_0_*, 0.092 in infectious period, 0.215 for *X_ss_* and 0.167 for *f_ss_*. Conversely, models trained and tested on trees with altered topologies (Dated Resolved-Model, Dated Polytomous-Model, Genetic Polytomous-Model and Genetic Resolved-Model) encountered challenges in accurately predicting superspreading parameters. This suggests that phylogenetic trees with polytomies lack sufficient phylogenetic resolution to accurately recover parameters related to superspreading. Models trained and tested on dated trees generally outperformed those trained and tested on the equivalent genetic distance trees in most scenarios, demonstrating the value of tip dates for informing model learning and estimating parameters.

**Fig. 3.**
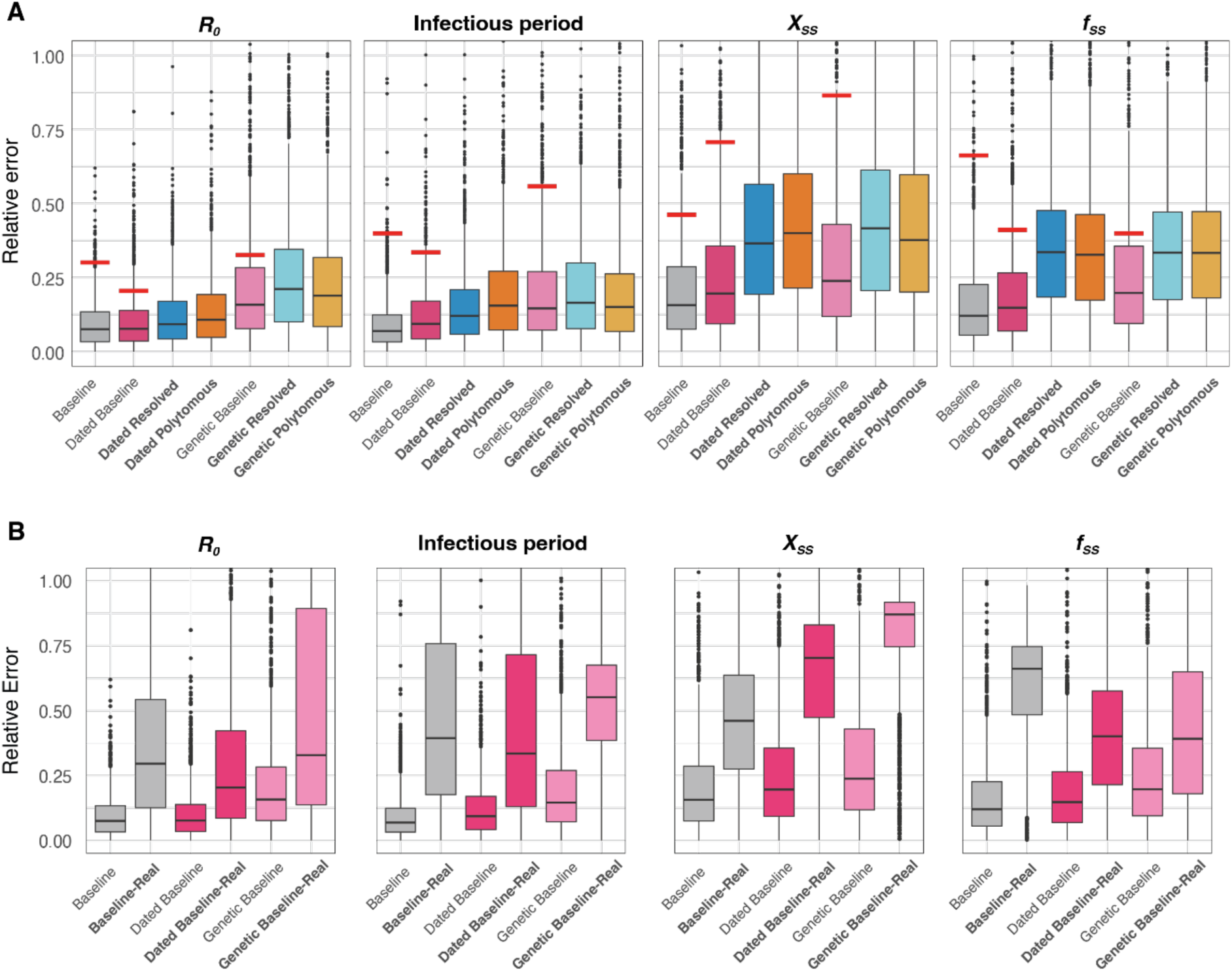
**Performance comparison of models.** A) Performance comparison of models trained on seven types of phylogenetic trees. Each bar depicts the relative error observed when testing trees of the same type as those used in training. The red marked lines denote the median relative error when testing the Baseline-Model and Dated Baseline-Model with Dated Resolved trees, as well as the Genetic Baseline-Model with Genetic Resolved trees. Models trained using realistic phylogenetic trees (i.e., Dated Resolved, Dated Polytomous, Genetic Resolved and Genetic Polytomous) are highlighted in bold. B) Performance comparison of models using realistic phylogenetic trees. “Baseline-Real” represents the evaluation of the Baseline-Model using Dated Resolved Trees. “Dated Baseline-Real” indicates the assessment of the Dated Baseline-Model with Dated Resolved Trees, while “Genetic Baseline-Real” reflects the performance of the Genetic Baseline-Model when utilizing Genetic Resolved trees.

### Impact of realistic phylogenetic trees on models trained with “ideal” trees

To evaluate the influence of using realistic phylogenetic trees as input on neural network models trained with “ideal” trees, we tested the Baseline-Model and Dated Baseline-Model with 1,000 Dated Resolved Trees and the Genetic Baseline-Model with 1,000 Genetic Resolved Trees (Fig.3 and Supplementary Table S1). The results revealed that the relative error for each parameter was approximately twice as high or more compared to when using “ideal” test trees. Notably, the relative errors for the superspreading parameters (*X_ss_* and *f_ss_*) were around or exceeded 0.5, and were worse than for models trained using realistic phylogenetic trees (such as Genetic Polytomous, Genetic Resolved, Dated Polytomous, and Dated Resolved) for both training and testing phases (Fig.3B). These findings suggest that PhyloDeep models trained on “ideal” trees struggle to predict accurately epidemiological parameters from realistic phylogenetic trees, but the accuracy of predictions can be improved when using models alternatively trained on more realistic trees. However, the higher predictive errors specific to superspreading parameters relative to other epidemiological parameters seemed to persist (Fig.3), highlighting the inherent challenge of estimating superspreading potential from phylogenetic trees. Additionally, despite repeatedly generating different Genetic Resolved and Dated Resolved trees from the polytomous trees as input, the predicted parameters tended to converge towards similar estimates, which differed substantially from the actual parameters originally input, thus indicating a form of bias in the estimations.

### Improving predictions by integrating contact tracing data

To improve model accuracy, a reasonable approach involves correcting the observed topology of input trees so that they closely resemble the equivalent “ideal” trees. With this context, we investigated the potential of leveraging contact tracing data to aid in refining the topology of Genetic Polytomous trees, for example, to match Baseline or Dated Baseline trees to varying extents (Supplementary Figure S4). We derived contact tracing information from the simulated Baseline trees, treating all descendants of each internal node as a cluster, with the dates of internal nodes considered as infection times of each cluster’s index case (Supplementary Figure S3). With this addition of cluster information and assuming perfect observation, the topology of Genetic Polytomous trees can be fully corrected (matching the genetic baseline trees), with external nodes subsequently dated to produce Dated Baseline trees (Supplementary Figure S4). Furthermore, if the infection times of clusters are known, time constraints can also be applied to internal nodes, effectively recovering equivalent Baseline trees from the Genetic Polytomous trees. In real-world scenarios, however, the extent of case observation is often limited and imperfect, and the accuracy of any available contact tracing data is uncertain and subject to additional biases.

Therefore, to assess how the quantity of contact tracing data influences our predictions within the context of phylogenetic trees, we simulated scenarios where 0, 25%, 50%, 75%, and 100% of internal nodes were randomly selected to provide cluster information and infection times. We then evaluated the performance of the Baseline-Model and Dated Baseline-Model (Fig. 4 and Supplementary Table S2). The former requires cluster information to resolve polytomies and infection times to estimate the lengths of newly created internal branches, while the latter relies solely on cluster information. For any remaining nodes lacking contact tracing data, we resolved them randomly as before. Our results indicated that even with just 25% of contact tracing data incorporated, the mean relative errors for *R_0_* and infectious time could be reduced to below 0.2, representing an improvement of 48% to 66% (Supplementary Table S2). As the availability of contact tracing data increased, model performance consistently improved, particularly in predicting superspreading parameters as could be expected. Incorporating 50% or more of contact tracing data yielded estimates of superspreading parameters, with mean relative errors around or below 30%, achieving an improvement of at least 22% (Supplementary Table S2). Notably, the Dated Baseline-Model generally outperformed the Baseline-Model except when contact tracing was 100% complete and a harsh time constraint margin of 0.1 day (Supplementary Table S2). Furthermore, the Dated Baseline-Model only required cluster information to refine the input trees, suggesting its greater relevance to real-world scenarios.

**Fig. 4.**
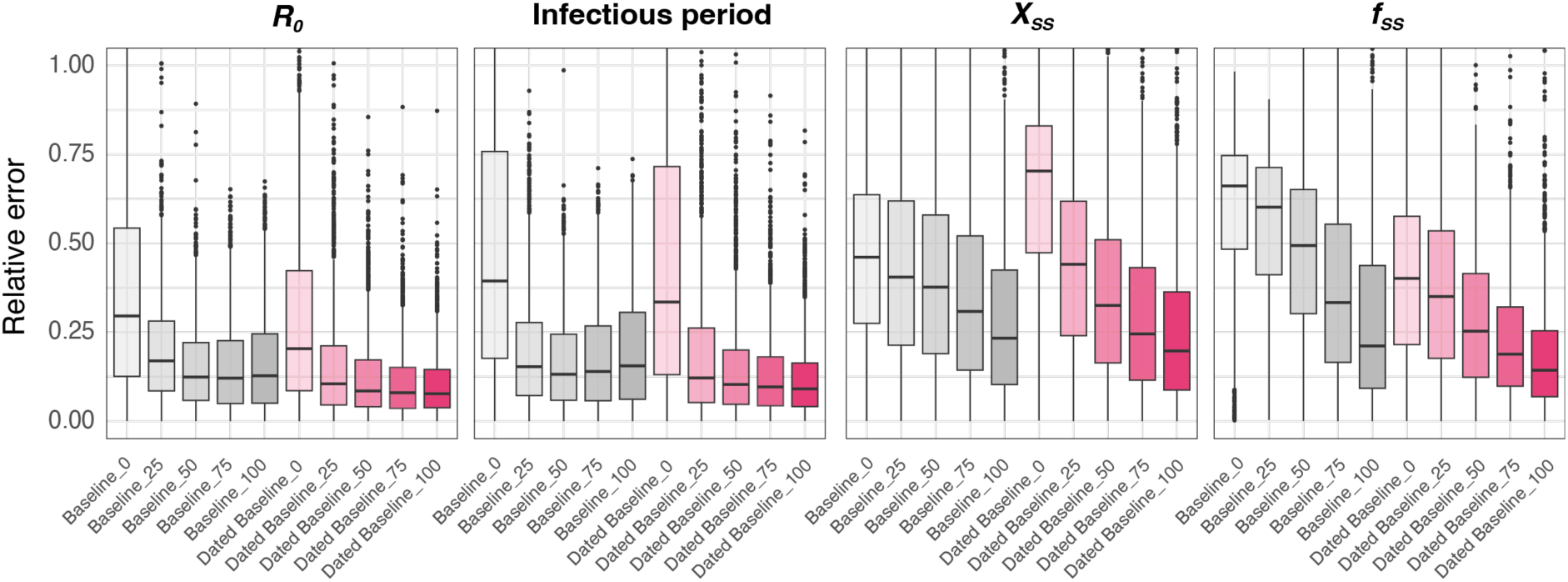
**Performance comparison by incorporating varying levels of contact tracing data based on Baseline-Model and Dated Baseline-Model.** The models are represented by grey (Baseline-Model) and red (Dated Baseline-Model) bars, with the color intensity within each bar signaling the degree of contact tracing data integrated into the input trees. Darker shades denote a higher percentage of data incorporation. The term “Baseline_50” refers to the performance of the Baseline-Model with Genetic Polytomous trees refined using 50% contact tracing data, encompassing cluster information and infection times. “Dated Baseline_50” indicates the performance of the Dated Baseline-Model with Genetic Polytomous trees refined using 50% contact tracing data, including cluster information.

### Case study of SARS-CoV-2 waves in Hong Kong

To demonstrate our method of integrating contact tracing data to improve model prediction, we used real-world SARS-CoV-2 data collected during the third and fourth waves of the epidemic in Hong Kong. By 2022, Hong Kong had effectively controlled the local spread of SARS-CoV-2, experiencing four significant waves during which extensive sequence sampling and epidemiological surveillance were conducted. Utilizing all available SARS-CoV-2 sequences from the third and fourth waves in Hong Kong, along with partial contact tracing data, we evaluate the differences in prediction outcomes when using the Dated Baseline-Model, with input trees refined by contact tracing data (Dated Resolved-Cluster) and without it (Dated Resolved).

Initially, we verified the suitability of the input trees through principal component analysis (PCA) and by comparing the range of each simulated SS to ensure the models and scenarios were predictive. All trees from Hong Kong passed this PCA check, but seven SSs related to superspreading features for the Dated Resolved tree of wave 4 were outside the [min, max] range of the simulated values (Supplementary Figure S1 and Table S3). After integrating the available contact tracing data (9.50%, as detailed in the Methods), only one SS remained outside the simulated range, albeit very close to the lower boundary (Supplementary Table S3).

The prediction results indicated a notable change when contact tracing data was used to refine tree topology, especially for wave 4 (Table 1). With the Dated Resolved-Cluster tree, we estimated an *R_0_* of 1.6 and 1.5, infectious periods of 4.6 and 8.6 days, *X_SS_* of 8.1 and 16.4, *f_ss_* of 0.09 and 0.08 for waves 3 and 4, respectively. Given *X_SS_* and *f_ss_*, we can calculate the dispersion value *k* (see Methods), which is commonly used as a measure of superspreading potential. For waves 3 and 4 we calculated *k* = 0.47 and 0.25 respectively, where lower values of *k* represent increasing superspreading potential. Conversely, using the Dated Resolved tree, we estimated an *R_0_* of 1.699 and 2.062, infectious periods of 5.720 and 20.071 days, *X_ss_* of 7.608 and 7.232, *f_ss_* of 0.090 and 0.076, and *k* of 0.488 and 0.658 for waves 3 and 4, respectively. Further, based solely on epidemiological records, we estimated an *R_0_* of 1.3 and 1.2, and *k* of 0.45 and 0.26 for waves 3 and 4, separately (Table 1). The observed discrepancies highlight the critical need for integrating diverse data sources and analytical methods in estimating epidemiological parameters, thereby enabling a more comprehensive and systematic understanding of epidemic dynamics.

**Table 1.** Comparison of inference of epidemiological parameters based on waves 3 and 4 of SARS-CoV-2 in Hong Kong.

| Waves | Input tree | $R_0$ | Infectious<br>period (day) | $X_{ss}$ | $f_{ss}$ | Dispersion<br>$k$ |
| --- | --- | --- | --- | --- | --- | --- |
| 3 | Dated Resolved | 1.699±0.096 | 5.720±1.018 | 7.608±1.496 | 0.090±0.022 | 0.488 |
|  |  | (1.460, 2.172) | (4.427, 10.804) | (4.141, 18.696) | (0.057, 0.163) | (0.441, 0.543) |
|  | Dated Resolved-Cluster | 1.588±0.077 | 4.636±0.635 | 8.078±1.709 | 0.091±0.021 | 0.467 |
|  |  | (1.330, 1.993) | (3.373, 8.238) | (3.911, 17.733) | (0.054, 0.167) | (0.418, 0.517) |
|  | Epidemiological<br>inference | 1.305 | NA | NA | NA | 0.451 |
|  |  | (1.146, 1.481) |  |  |  | (0.421, 0.481) |
| 4 | Dated Resolved | 2.062±0.072 | 20.071±1.663 | 7.232±1.423 | 0.076±0.009 | 0.658 |
|  |  | (1.628, 3.220) | (14.235, 32.668) | (2.197, 23.198) | (0.050, 0.154) | (0.596, 0.737) |
|  | Dated Resolved-Cluster | 1.518±0.091 | 8.629±0.881 | 16.388±2.692 | 0.078±0.007 | 0.250 |
|  |  | (1.284, 2.055) | (6.548, 14.929) | (5.895, 33.409) | (0.050, 0.161) | (0.227, 0.278) |
|  | Epidemiological<br>inference | 1.212 | NA | NA | NA | 0.264 |
|  |  | (1.042, 1.406) |  |  |  | (0.248, 0.279) |

Note: Values predicted by neural network models are expressed as mean ± standard deviation generated by randomly resolving poytomies $n = 200$ times. Values in parentheses are the 95% CI.

Additionally, we conducted 200 random resolutions of polytomies for these SARS-CoV-2 trees to measure the robustness of the predictions. The resulting standard deviation were notably small (Table 1), indicating that the predictions were not significantly affected by the random resolution of polytomies, suggesting our models could efficiently extract essential cluster information and guide robust predictions. The 95% confidence intervals (CIs) were generated by parametric bootstrap as per the methodology of PhyloDeep. The substantial width of CIs for superspreading parameters again highlight the inherent difficulty in predicting these metrics.

## Discussion

In this study, we assessed the performance of established neural network models (PhyloDeep) in predicting epidemiological parameters and the applicability of these models to real-world scenarios using SARS-CoV-2 as a case study for both simulation and empirical analyses. Our findings demonstrate the relative performance limitations of utilizing neural network models trained on simulated phylogenetic trees (“ideal” trees) when predicting parameters from real-world trees featuring polytomies, and show that models alternatively trained on more realistic trees can improve the accuracy of predictions. Beyond upstream improvements to model training, we show that by using contact tracing data to partially adjust the topology of input trees downstream, additional performance enhancements can be achieved. We apply this approach to SARS-CoV-2 genome sequences from Hong Kong matched to minimal contact tracing data, producing new phylodynamic estimates of both *R* and *k*.

Without the incorporation of contact tracing data, we found that even our improved models trained on more realistic trees struggled to accurately estimate parameters related to superspreading. This issue is particularly pronounced when sequences are nearly identical, like for SARS-CoV-2, which results in potentially biased clustering likely to misinform public health decision makers. Traditional phylodynamic models (e.g. maximum likelihood estimation and Bayesian approaches), which assume ideal binary trees and not representing sequence evolution, also struggle in parameter estimation under these conditions. Together this emphasizes the importance of incorporating even minimal contact tracing data as we have done in our study, but also utilizing more comprehensive summary statistics focused on clusters or polytomies that can effectively capture the complexity of the underlying transmission dynamic. One previous study (Tran-Kiem & Bedford, 2024) has demonstrated a connection between the size distribution of identical sequence clusters and transmission dynamics, however, our attempts to incorporate similar information into our neural network models, trained on genetic distance trees, yielded limited improvements. As an ongoing area of research interest, future study could evaluate the relative predictive performance of models that expand the potential range of features related to clusters or polytomies.

Besides superspreading, the incubation period is another significant aspect of pathogen transmission dynamics. For example, estimates of the SARS-CoV-2 incubation period were used to justify the World Health Organization’s (WHO) recommendation of a 14-day quarantine period for contacts of infected cases (Wells et al., 2021). In our approach, we utilized a BDSS model, which does not account for the incubation period, but defines the infectious period as the interval from infection time to sampling date otherwise known as the delay interval. Employing the Dated Baseline-Model with the Dated Resolved-Cluster tree, we determined that the infectious period/delay interval of waves 3 and 4 to be approximately one week, however the delay for wave 4 was longer than that for wave 3, suggesting case detection speed was somewhat challenged. The longer delay in wave 4 could be explained by the sudden rise in cases associated with the largest single SARS-CoV-2 superspreading event detected in Hong Kong prior to widespread vaccination, which also triggered the start of wave 4. The clear bimodal epidemic observed in wave 4, compared to the classic unimodal pattern observed in wave 3, also affirms that case detection speed may have suffered at that time, in line with our estimates (Adam et al., 2022).

Remarkably, the estimation of *R_0_* exhibited robust performance across our neural network models, with models trained on dated trees outperforming those based on genetic distance trees. This underscores the value of tip dates for *R_0_* estimation, particularly as sequence variability decreases. This is in line with recent studies that highlight the increasing importance of sampling dates for phylodynamic inference when sequence variability is low (Featherstone et al., 2023). When realistic trees were used as input, models like the Dated Resolved-Model and Dated Polytomous-Model showed excellent performance, suggesting their potential for effective and accurate *R_0_* and infectious period predictions from sequence data. This offers a promising avenue for tracking epidemic dynamics using sequence data, which, when compared with epidemiological records, can provide deeper insights and mitigate potential sampling biases. Future investigations are needed to ascertain the extent to which sequence data can facilitate robust predictions and to evaluate the effects of progressively incorporating new sequence samples.

Our study acknowledges certain limitations. Notably, the BDSS model does not account for the incubation period of the disease, introducing a significant source of uncertainty. The omission of the incubation period from our transmission models necessitates further exploration in future studies to mitigate these uncertainties. Additionally, real-world contact tracing data may contain inherent biases or inaccuracies. In applying our model to the SARS-CoV-2 dataset from Hong Kong, we presumed the accuracy of the contact tracing data. This assumption allowed us to collapse all associated children (see Methods), including those are not recorded within the cluster, potentially leading to an inaccurate refinement of the tree topology and biased predictions.

Importantly, making trees realistic hinges on the specific sequence length and evolution rate of SARS-CoV-2, rendering these models in this study inapplicable to other viruses. To extend their use to other pathogens, modifications are required to accommodate variations in sequence length and evolution rate, building pathogen-specific models as we show for SARS-CoV-2. This contrasts with PhyloDeep, which is adaptable for studying a diverse array of pathogens. Correspondingly, the choice of a specific birth-death model emerges as another crucial factor that must be carefully considered.

Overall, this study highlights the challenges of relying solely on viral phylogenetic trees generated from sequences for estimating superspreading events. The integration of even minimal contact tracing data can significantly enhance model predictions, emphasizing the importance of such data in surveillance efforts for emerging infectious diseases, particularly when viral sequences lack variability. We hope our comprehensive evaluation will inform future developments in deep learning applications within phylogenetics and phylodynamics.

## Methods

### Simulations

In this study, SARS-CoV-2 served as the reference pathogen for evaluating the performance of the existing deep learning model PhyloDeep. Given the marked overdispersion in SARS-CoV-2 transmission dynamics, characterized by superspreading (Adam et al., 2020; Du et al., 2022; Guo et al., 2022) we used treesimulator (v0.1.7: https://github.com/evolbioinfo/treesimulator/releases/tag/0.1.7) to generate time-scaled phylogenetic trees. These trees were generated with a BDSS model, distinguishing cases into superspreaders (S) and normal spreaders (N), in addition to the conventional parameterisation of the Birth-Death model, i.e. *R_0_* and the infectious period. Superspreaders constitute a small fraction of the total simulated population (denoted by *f_SS_* = *β_SS_*/*β_SS_* + *β_SN_*)) but can transmit the virus at rates significantly higher than normal spreaders, where superspreading transmission rates are denoted as *X_SS_* = *β_SS_*/*β_NS_* = *β_SN_*/*β_NN_*). Upon reviewing the 98 summary statistics (SS) (see details in Feature representation and neural network models section), it was noted that certain metrics associated with branch lengths and superspreading events based on the SARS-CoV-2 dataset from Hong Kong fell outside the [min, max] range of simulated values in PhyloDeep, characterized by a lower median/mean SS and increased variance SS (detailed in Supplementary Table S4). Consequently, to better capture the complexities of SARS-CoV-2 transmission dynamics, we expanded the range of epidemiological parameters for tree simulation in PhyloDeep, summarized in Supplementary Table S5.

Simulated time-scaled trees are transformed into Genetic Baseline trees, with branch lengths determined by a binomial process, *B* (*n*=sequence length, *p*=evolutionary rate × branch length of time-scaled trees). For SARS-CoV-2, the sequence length is 29,903, and the evolutionary rate has a mean of 8×10^−4^ and a standard deviation of 4×10^−4^ substitutions per site per year, with a lognormal distribution (Hadfield et al., 2018; Jolly & Scaria, 2021). In Genetic Baseline trees, branches representing zero mutation are collapsed to form Genetic Polytomous Trees. Within these trees, polytomies are resolved by randomly coalescing two offspring until binary trees, termed Genetic Resolved Trees, are obtained. These genetic distances are then re-dated using LSD2 (To et al., 2016), assigning dates to the tips by adding the lengths from the tips to the root within the time-scaled trees to a dummy date designated as the root date. Additionally, a temporal constraint for the root is established by setting a range (dummy date − 1 day, dummy date + 1 day), ensuring the root’s time is not excessively early. The clock rate used is the same as mentioned above, with a mean of 8×10^−4^ and a standard deviation of 4×10^−4^ substitutions per site per year.

An additional 100,000 trees were simulated, and the PhyloDeep methodology was applied to establish the 95% CIs.

### Feature representation and neural network models

We represent time-scaled phylogenetic trees using sampling probability and 98 SS, as employed in PhyloDeep (Saulnier et al., 2017; Voznica et al., 2022). However, for genetic distance trees, certain concepts like transmission chains (14 SS) associated with superspreading and lineage through time (LTT) (49 SS) are not directly applicable. To address this, we designed 62 SS focused on the distribution of internal (31 SS) and external (31 SS) nodes by counting the nodes that are n (0-30) mutations away from the tree root. Additionally, the size distribution of clusters of identical sequences, which is indicative of transmission dynamics and heterogeneity (Tran-Kiem & Bedford, 2024), led us to include 10 SS related to the distribution of identical sequence clusters, with sizes ranging from 1 to 10. Consequently, 90 SS are utilized to characterize the genetic distance tree. While time-scaled trees are rescaled so the average branch length equals 1 prior to representation (Voznica et al., 2022), genetic distance trees do not require this adjustment.

Following the PhyloDeep methodology, we developed our neural network model using Python 3.6, with the Tensorflow 1.5.0, Keras 2.2.4, and scikit-learn 0.19.1 libraries. The model comprises an input layer with 99 or 90 nodes, four sequential hidden layers arranged in a funnel shape with 64, 32, 16, and 8 neurons, respectively, and an output layer that predicts four parameters: R0, infectious period, *X_ss_*, and *f_ss_*. The neurons in the last hidden layer utilize linear activation, whereas the others employ exponential linear (ELU) activation. The performance of our neural network models is assessed as the mean relative error (MRE) of the estimator:

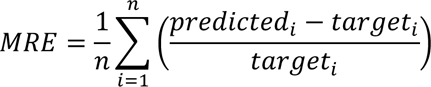

where *n* is the number of simulated trees used in the test set.

To draw a parallel with epidemiological inference, *X_ss_* and *f_ss_* can be transformed into the dispersion *k*. Utilizing the multi-type birth-death model process (Stadler & Bonhoeffer, 2013), it becomes possible to estimate the probability of an individual infecting “n” others over its lifespan, aligning with a geometric distribution. By synthesizing the probability with the cumulative number of infections, the offspring distribution was ascertained. The approach outlined in “Estimating *R_0_* and *k* from epidemiological data only” section was employed to derive *k* from this offspring distribution.

### Integration of contact tracing data into phylogenetic trees

In our simulations, we utilize time-scaled trees to derive contact tracing data, treating all descendants of each internal node as a single cluster, with the node’s age representing the infection time (Supplementary Figure S3). Using such contact tracing data, we refine the phylogenetic trees by identifying the most recent common ancestor (MRCA) for each cluster. We then iterate through children of the MRCA and coalesce all associated children, encompassing both leaves and children of internal nodes within the cluster. This process enables us to resolve polytomies in Genetic Polytomous trees, facilitating their transformation back into Genetic Baseline trees (Supplementary Figure S4).

Additionally, by applying the infection times as time constraints on the internal nodes, we can revert Genetic Baseline trees to their Baseline counterparts using LSD2 (To et al., 2016). We achieve this by setting a specific time range for the internal nodes, using a margin of (infection time − 1 day, infection time + 1 day). Narrowing this margin to 0.1 day brings the converted trees even closer to the Baseline trees, thereby yielding performance on the Baseline-Model that is nearly identical to that obtained when directly using Baseline trees for testing, as detailed in Supplementary Tables S1 and S2.

### SARS-CoV-2 dataset in Hong Kong

We used sequences and epidemiological data from the third and fourth waves of SARS-CoV-2 in Hong Kong, as detailed in our prior study (Gu et al., 2022). These waves were characterized by single introduction events that sparked local transmissions, and they were notable for their relatively consistent sequence sampling and comprehensive surveillance data. In this study, we focused on the exponential stages of waves 3 and 4, which spanned from May 13 to August 1, 2020, with 460 sequences and 1,930 local cases, and from September 30 to December 8, 2020, with 243 sequences and 1,577 local cases, respectively. The sampling rates for waves 3 and 4 were 23.8% and 15.4%, respectively. During wave 3, 84.35% (388 out of 460) of sequences were linked to cluster information involving 191 clusters, among which 76 clusters comprised more than one sequence. This indicates that 16.56% (76 out of 459) of the data were supported by contact tracing. In wave 4, 90.53% (220 out of 243) of sequences were associated with 35 clusters, with 23 clusters containing multiple sequences, amounting to 9.50% (23 out of 242) contact tracing data availability.

For waves 3 and 4, we reconstructed Maximum Likelihood (ML) phylogenies using RAxML-NG (Kozlov et al., 2019) with the GTR+G4+FO substitution model. We maintained consistency with simulated trees in terms of collapsing internal nodes and the random resolution of polytomies. Our findings revealed that the distribution of the number of offspring from collapsed internal nodes falls within the range observed in our simulations (Supplementary Figure S2). Subsequently, these trees were dated using LSD2 (To et al., 2016), following a strict molecular clock assumption of 8×10^−4^ substitutions per site per year (Hadfield et al., 2018; Jolly & Scaria, 2021), and applying time constraints for the root as inferred by (Gu et al., 2022).

### Estimating *R_0_* and *k* from epidemiological data only

We compared the results for *R_0_* and *k* estimated using PhyloDeep to those estimated with the same epidemiological data on SARS-CoV-2 available during the exponential periods of waves 3 and 4 in Hong Kong. Epidemiological estimates of *R_0_* were generated using EpiNow2 (Sam Abbott, 2020) and case reports, where *R_0_* was calculated as the mean and 95% quantiles of *R_t_* estimated during waves 3 (May 13 to August 1, 2020) and wave 4 (September 30 to December 8, 2020). *R_t_* was estimated using an empirical delay distribution of symptomatic SARS-CoV-2 cases in Hong Kong, calculated as the difference in days between symptom onset and report dates, excluding negative delays where cases were reported prior to the recorded symptom onset date. Additionally, we used an uncertain gamma distribution for the incubation period (Lauer et al., 2020) (mean = 3.6, mean SD = 0.71, and SD = 3.1, SD SD = 0.77) and uncertain lognormal distribution for the generation time (Ganyani et al., 2020) (logmean = 1.6, logmean SD = 0.064, and logSD = 0.42, logSD SD = 0.069) to estimate *R_t_* at the time of infection.

Epidemiological estimates of *k* estimate were generated from empirical offspring distributions for SARS-CoV-2 available from previous studies in Hong Kong (Adam et al., 2022). These distributions were generated from infector-infectee pairs, where the number of secondary cases is counted for each unique infector and includes chain-terminating infectees as zero. We subsetted the empirical offspring distributions to the same exponential periods for wave 3 and wave 4 as before, given the estimated infection date of each paired case as a deconvolution of the generation time, incubation period, and delay distributions given the onset date or report dates if asymptomatic between infector-infectee pairs. Importantly, offspring counts were not artificially right-censored, meaning the observed count of each infector case was included even if the estimated infection date of paired infectee(s) fell after the exponential periods of each wave. Following the approach of Llyod-Smith et al (Lloyd-Smith et al., 2005), we directly estimate *k* from the finalised offspring distributions by maximum likelihood estimation, assuming a negative binomial model jointly parameterised by the mean and dispersion parameter *k,* with 95% intervals generated by non-parametric bootstrap estimation sampling 1000 replicates with replacement.

## Data Availability

All data produced in the present work are contained in the manuscript

## Acknowledgments

We acknowledge the technical support provided by colleagues from the Centre for PanorOmic Sciences of the University of Hong Kong. We also acknowledge the Centre for Health Protection of the Department of Health for providing epidemiological data for the study. The computations were performed using research computing facilities offered by Information Technology Services, the University of Hong Kong. The funding bodies had no role in the design of the study and collection, analysis, and interpretation of data and writing of the manuscript.

## Data and materials availability

All anonymized data, code, and analysis files are available in the GitHub repository (https://github.com/vjlab/dl-phylodynamics-ct).

