## Supplementary materials for "Deep learning of SARS-CoV-2 outbreak phylodynamics with contact tracing data"

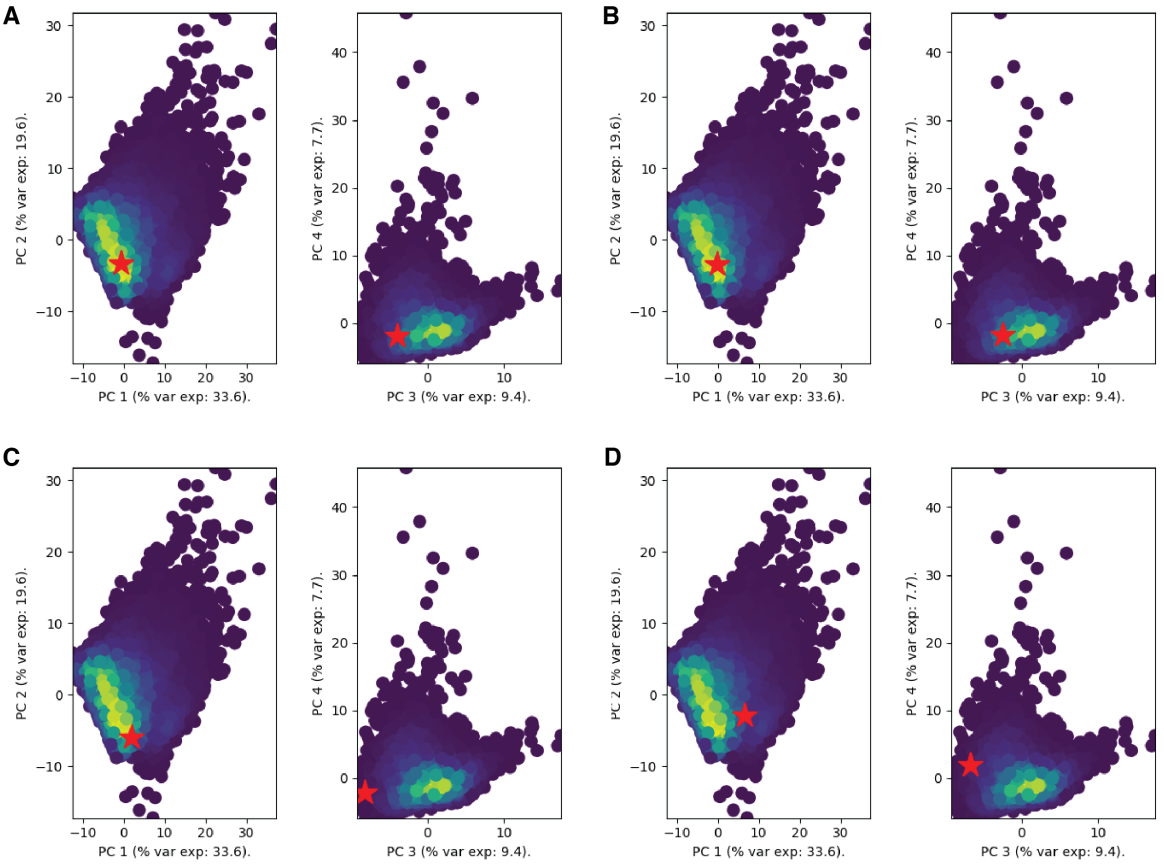


Supplementary Figure 1 | PCA plots illustrating the third and fourth waves of SARS-CoV-2 in Hong Kong. For the third wave: Dated Resolved (A) and Dated Resolved-Cluster (B). For the fourth wave: Dated Resolved (C) and Dated Resolved-Cluster (D).

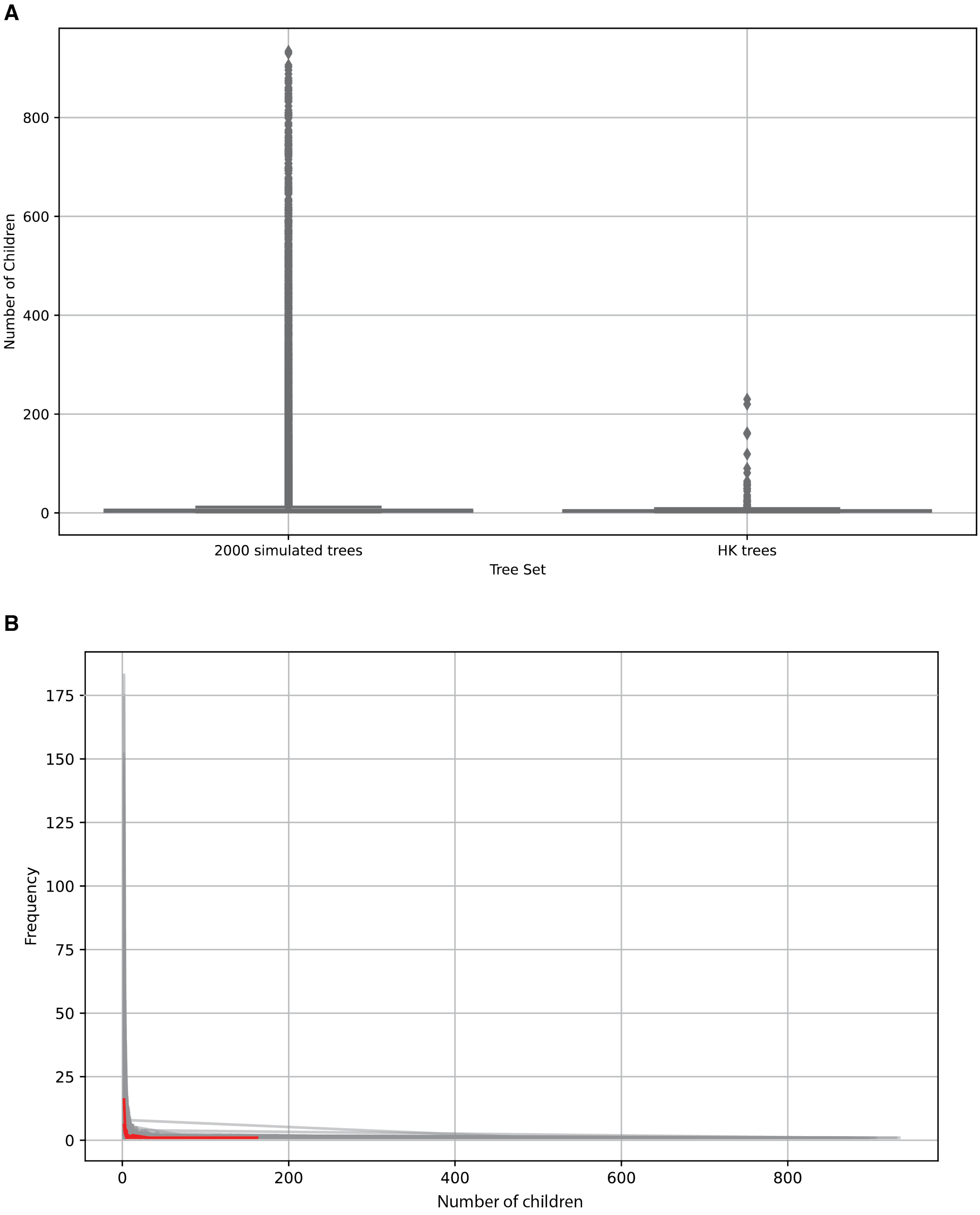


**Supplementary Figure 2** | A comparison of the distribution of internal node children between 2000 simulated trees and trees generated from SARS-CoV-2 sequences during waves 3 and 4 in Hong Kong. (A) Contrasts children per internal node in simulated trees (left) with those observed in SARS-CoV-2 trees ( right). (B) Depicts internal node offspring frequency across trees, with each line representing one simulated tree. 3rd and 4th wave occurrences are highlighted in red.

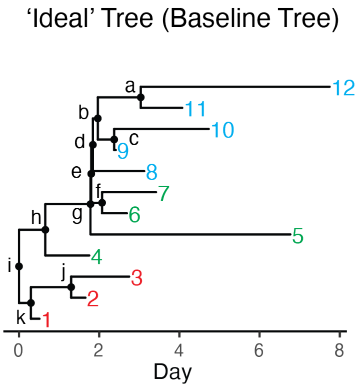


Contact tracing data

| *Internal nodes* | *Cluster information* | *Infection time* |
|---|---|---|
| a | 11,12 | TMRCA(11,12) |
| b | 9,10,11,12 | TMRCA(9,10,11,12) |
| c | 9,10 | TMRCA(9,10) |
| d | 8,9,10,11,12 | TMRCA(8,9,10,11,12) |
| e | 6,7,8,9,10,11,12 | TMRCA(6,7,8,9,10,11,12) |
| f | 6,7 | TMRCA(6,7) |
| g | 5,6,7,8,9,10,11,12 | TMRCA(5,6,7,8,9,10,11,12) |
| h | 4,5,6,7,8,9,10,11,12 | TMRCA(4,5,6,7,8,9,10,11,12) |
| i | 1,2,3,4,5,6,7,8,9,10,11,12 | TMRCA(1,2,3,4,5,6,7,8,9,10,11,12) |
| j | 2,3 | TMRCA(2,3) |
| k | 1,2,3 | TMRCA(1,2,3) |

**Supplementary Figure 3** | Extraction of contact tracing data from an “ideal” tree.

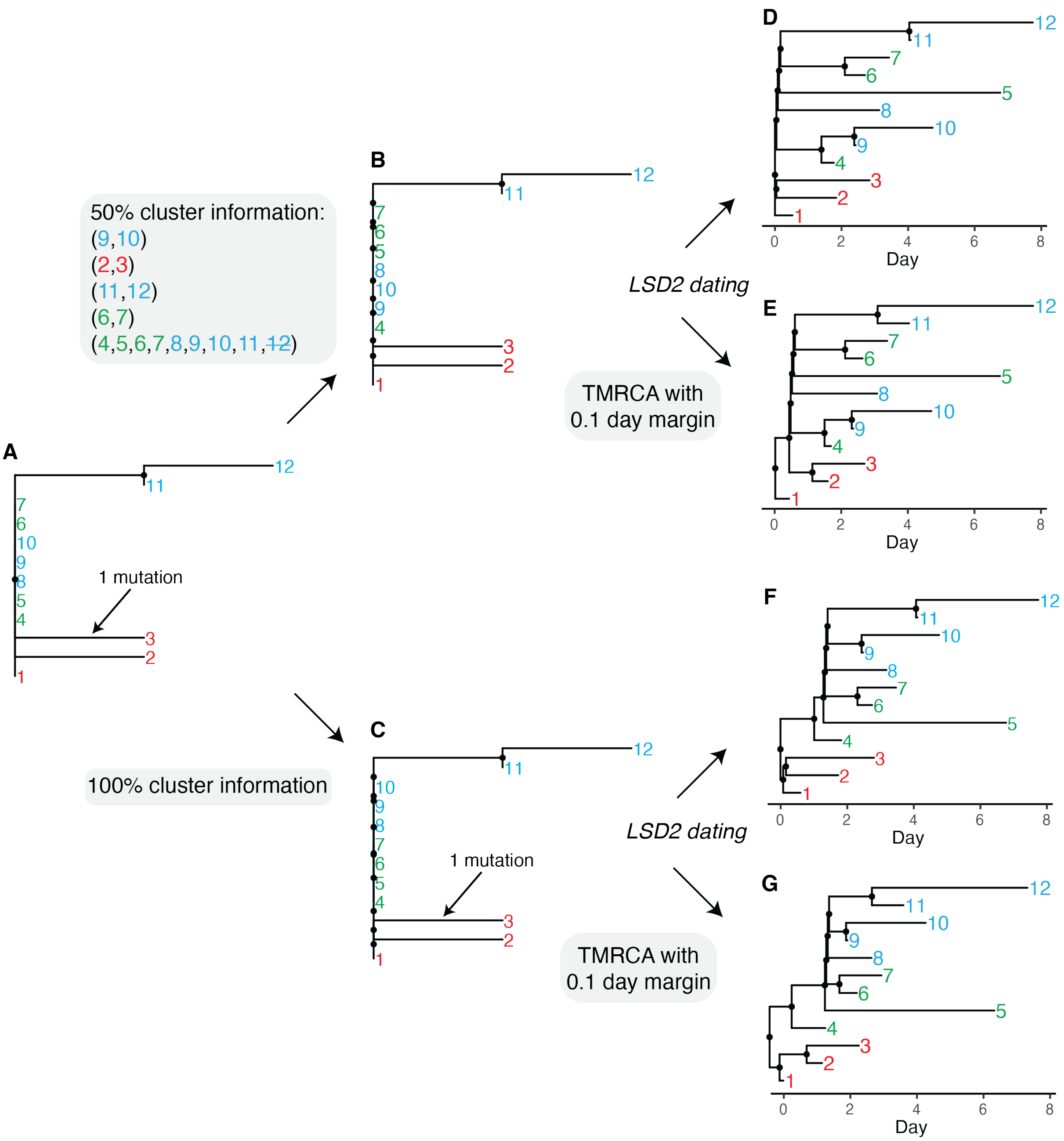


**Supplementary Figure 4** | Resolving polytomies in the Genetic Tree using contact tracing data. (A) The initial Genetic Polytomous Tree, as in **Fig. 1**. (B) Tree resolved with 50% cluster information; notably, the inclusion of sample '12' in the fifth cluster does not alter the resolution outcome per our methodology. Polytomies lacking cluster information were randomly resolved into a binary tree. (C) Fully resolved tree with 100% cluster information, matching the Genetic Baseline Tree in **Fig. 1**. (D, E) Dated trees from tree in Panel (B) using LSD2, without and with temporal constraints from known 50% clusters, respectively. (F) Tree from Panel (C) dated using LSD2, corresponding to the Dated Baseline Tree in **Fig. 1**. (G)

Tree from Panel (C) dated using LSD2 with temporal constraints applied to all clusters, closely resembling the "ideal" tree in **Fig. 1**.

**Supplementary Table 1** | Performance comparison (mean relative error) of models trained on seven types of phylogenetic trees.

| Model | $R_0$ | *Infectious period* | $X_{ss}$ | $f_{ss}$ |
|---|---|---|---|---|
| Baseline-Model | 0.095 | 0.092 | 0.215 | 0.167 |
| | (0.382)* | (0.561) | (0.721) | (0.598) |
| Dated Baseline-Model | 0.103 | 0.122 | 0.268 | 0.200 |
| | (0.306) | (0.566) | (0.648) | (0.465) |
| Dated Polytomous-Model | 0.136 | 0.196 | 0.449 | 0.357 |
| Dated Resolved-Model | 0.119 | 0.155 | 0.444 | 0.367 |
| Genetic Baseline-Model | 0.205 | 0.191 | 0.314 | 0.265 |
| | (0.664) | (0.523) | (0.798) | (0.602) |
| Genetic Polytomous-Model | 0.232 | 0.192 | 0.454 | 0.361 |
| Genetic Resolved-Model | 0.261 | 0.214 | 0.480 | 0.360 |

**Values in parentheses represent performance metrics tested with realistic phylogenetic trees: the Baseline-Model and Dated Baseline-Model analyses utilized Dated Resolved trees, whereas the Genetic Baseline-Model employed Genetic Resolved trees for evaluation.*

**Supplementary Table 2** | Performance comparison (mean relative error) by incorporating varying levels of contact tracing data based on Baseline-Model and Dated Baseline-Model.

| Model | *Percentage of contact tracing data* | $R_0$ | *Infectious period* | $X_{ss}$ | $f_{ss}$ |
|---|---|---|---|---|---|
| Baseline-Model | 0% | 0.382 | 0.561 | 0.721 | 0.598 |
| | 25% | 0.198 | 0.193 | 0.667 | 0.544 |
| | 50% | 0.154 | 0.165 | 0.524 | 0.467 |
| | 75% | 0.154 | 0.178 | 0.395 | 0.370 |
| | 100% | 0.165 (0.101)* | 0.196 (0.109) | 0.302 (0.222) | 0.308 (0.194) |
| Dated Baseline-Model | 0% | 0.306 | 0.566 | 0.648 | 0.465 |
| | 25% | 0.156 | 0.200 | 0.485 | 0.364 |
| | 50% | 0.124 | 0.150 | 0.392 | 0.285 |
| | 75% | 0.111 | 0.132 | 0.323 | 0.229 |
| | 100% | 0.103 | 0.120 | 0.266 | 0.187 |

**Values in parentheses represent performance metrics tested with a time constraint margin of 0.1 day.*

**Supplementary Table 3** | Summary of out-of-range SS for waves 3 and 4 compared to simulated values in Dated Baseline-Model.

| Features | Max | Min | *Dated Resolved (wave 3)* | *Dated Resolved-Cluster (wave 3)* | *Dated Resolved (wave 4)* | *Dated Resolved-Cluster (wave 4)* |
|---|---|---|---|---|---|---|
| Mean length of transmission chain | 2.979 | 0.548 | 0.643 | 0.676 | **0.247*** | **0.452** |
| The 4th deciles of transmission chain | 2.155 | 0.019 | 0.050 | 0.053 | **0.015** | 0.029 |
| The 5th deciles of transmission chain | 2.552 | 0.028 | 0.050 | 0.053 | **0.015** | 0.029 |
| The 6th deciles of transmission chain | 2.948 | 0.043 | 0.050 | 0.288 | **0.015** | 0.177 |
| The 7th deciles of transmission chain | 3.778 | 0.065 | 0.156 | 0.559 | **0.015** | 0.456 |
| The 8th deciles of transmission chain | 4.789 | 0.102 | 0.651 | 0.883 | **0.016** | 0.621 |
| Max length of transmission chain | 29.365 | 2.146 | 5.372 | 4.453 | **2.115** | 3.270 |

**Values highlighted in bold exceed the established maximum and minimum range thresholds.*

**Supplementary Table 4** | Summary of out-of-range SS for waves 3 and 4 compared to simulated values in PhyloDeep.

| **Features** | **Max** | **Min** | ***Dated Resolved (wave 3)*** | ***Dated Resolved (wave 4)*** |
|---|---|---|---|---|
| Median of all branch length | 0.939 | 0.520 | **0.445*** | **0.402** |
| Variance of all branch length | 1.636 | 0.590 | **1.737** | **1.886** |
| Variance of external branch length | 2.095 | 0.363 | **2.288** | **2.622** |
| Mean length of transmission chain | 2.481 | 0.929 | **0.643** | **0.247** |
| The 2nd deciles of transmission chain | 1.382 | 0.186 | **0.050** | **0.015** |
| The 3rd deciles of transmission chain | 1.663 | 0.298 | **0.050** | **0.015** |
| The 4th deciles of transmission chain | 1.867 | 0.411 | **0.050** | **0.015** |
| The 5th deciles of transmission chain | 2.063 | 0.522 | **0.050** | **0.015** |
| The 6th deciles of transmission chain | 2.257 | 0.627 | **0.050** | **0.015** |
| The 7th deciles of transmission chain | 2.672 | 0.775 | **0.156** | **0.015** |
| The 8th deciles of transmission chain | 3.178 | 0.862 | **0.651** | **0.016** |
| The 9th deciles of transmission chain | 3.635 | 1.214 | 1.257 | **0.345** |
| The 10th deciles of transmission chain | 5.241 | 1.419 | 2.071 | **0.859** |
| Max length of transmission chain | 15.296 | 2.189 | 5.372 | **2.115** |

**Values highlighted in bold exceed the established maximum and minimum range thresholds.*

**Supplementary Table 5** | Parameter ranges used for simulations.

| Parameters | Range in PhyloDeep | Range in this study |
|---|---|---|
| $R_0$ | (1, 5) | (**0.4**, **6**)* |
| Infectious period | (1, 10) | (1, **14**) |
| $X_{ss}$ | (3, 10) | (**2**, **40**) |
| $f_{ss}$ | (0.05, 0.20) | (0.05, **0.25**) |
| $k$ | (0.33, 0.90) | (**0.09**, **1.18**) |
| *Tips* | (200, 500) | (200, **1000**) |

**Values highlighted in bold indicate the adjusted boundary values.*
